# Impact of Emerging COVID-19 variants on psychosocial health: A Systematic Review

**DOI:** 10.1101/2023.07.23.23293040

**Authors:** Pratyush Kumar, Manali Sarkar, Morales Femenias Yurkina, Ramya Gnanaraj, Daniel Jesús García Martínez, Yhojar A. Pisfil-Farroñay, Laxmi Chaudhary, Poonam Agrawal, G. P. Kaushal, Mathew Mbwogge, Kumar Abhishek, Muhannad Alnaasan, Maximiliano Ezequiel Arlettaz, Reem Kozum, Miguel Fernando Juárez Moyrón, Suhrud Panchawagh, Asmitha P Reddy, Vishnu B Unnithan, Rushikesh Shukla

## Abstract

**Background:** The COVID-19 pandemic has had significant psychological effects on individuals and communities around the world. Studies have found that the prevalence of anxiety and depression symptoms increased significantly during the pandemic. The goal of the study is to understand how the emerging new virus variants keep the world in a state of fear and the ways in which mental health measures can be implemented and adopted to alleviate anxiety.

**Methods:** A broad search for observational studies were carried out in Pubmed, Google Scholar, Clinical Key, and World Medical Library. Studies that reported and/or related the existence of anxiety generated by suffering or not from diseases caused by the new emerging Covid-19 viruses and that for which the full text of the article was accessible were included in the study while systematic review and meta-analysis and studies in groups were excluded.

**Results:** 22 studies were included in the review. The deleterious psychosocial effects were the restructuring of life, establishment of unhealthy habits, emergence of “*corona phobia*”, fear and stigma of being afflicted with the disease and spreading it to loved ones, and lack of contact with others. Increased rates of depression and anxiety were also seen. The circulating variants responsible for these main psychosocial repercussions were: *Epsilon, Zeta, Eta, Iota, Kappa, Alpha, Beta, Gamma, and Delta*. Social support was found to be protective.

**Conclusion:** Hence interventions targeted at promoting mental health should be considered a public health priority.

## INTRODUCTION

The COVID-19 pandemic has had significant psychological effects on individuals and communities around the world. The fear and uncertainty surrounding the virus [1] as well as the loss of loved ones [2], job insecurities [3], financial hardships [4], reduced sleep quality [5], and forced social isolation caused by lockdown measures [6] have all contributed to mental health challenges for many people.

One of the main psychological effects of the pandemic has been an increase in stress and anxiety. Studies have found that the prevalence of anxiety and depression symptoms increased significantly during the pandemic. [7,8] Other researchers have found that the pandemic has disproportionately affected vulnerable populations, such as those with pre-existing mental health conditions [9] and those who have experienced trauma or adversity. The risk of suicidal ideation increased, especially among adult. [10] The pandemic affected all age groups including children, with school closures [11], adolescents and women due to social isolation, withdrawal from social life [12] and due to domestic violence[13]. The pandemic has been particularly difficult for older adults, who may be more isolated to begin with and may be more vulnerable to the negative effects of loneliness [14].

COVID also affected people from all walks of life, including healthcare workers [15], remote workers of the information system [16], small-scale business owners and entrepreneurs [17] migrant workers [18], etc., thereby causing economic turndown and recession which in turn led to unemployment, layoffs [19] and financial hardships leading to increased stress and anxiety. Additionally, the world-recognized worry and anxiety brought on by the pandemic have recently challenged and replaced psychological well-being, which has long been acknowledged as the mental health benefit of travel [20].

The emergence of COVID-19 variants, including the Delta, Gamma variants and the Omicron variants, and the vaccine variants [21] has added to this stress and anxiety. The uncertainty surrounding the spread and impact of these variants, as well as the potential for additional lockdowns and other measures to control their spread, has caused increased worry and concern for many people with an additional layer of fear of re-infection and the potential for more severe illness becoming a reality. The goal of the study is to understand how the emerging new virus variants keep the world in a state of fear and the ways in which mental health measures can be implemented and adopted to alleviate anxiety.

## METHODOLOGY

### Search strategy

A broad search for observational studies was carried out, which evaluated the repercussions or psychological and social consequences in the child and adult population during the outbreak of SARS-COV 2 and its new variants. The databases used during the search were: Pubmed, Google scholar, Clinical Key and World Medical Library. The Boolean operators used for the search strategy in the databases were: Coronavirus OR quarantine AND psychological effects OR psychosocial OR distress OR mental health OR mental health care, and their equivalents in Spanish, in title and abstract, in published scientific articles. between March 2020 and May 2022. As exclusion criteria, the following were considered: studies in groups other than the child and adult population, examples of studies of students, of health personnel.

We searched for all randomized and non-randomized studies that reported the existence of anxiety in the face of new emerging variants of the virus (Covid-19), in both the child and adult population and that were published between March 2020 and May 2022; For this, the databases were used: CENTRAL, MEDLINE through PubMed and Embase. In addition, a search was carried out in the citation indexes: Google scholar, Scopus and World Medical Library; this was done to screen for possible further studies.

The search in CENTRAL, Pubmed and Embase was: (covid or covid-19 OR coronavirus OR “coronavirus” OR SARSCoV-2 OR “Coronavirus”[Mesh] OR “severe acute respiratory syndrome coronavirus 2”[Complementary concept] OR “COVID-19”[Supplementary concept] Concept] OR “Coronavirus infections”[Mesh] OR “Coronavirus infections/psychology”[Mesh] OR “Coronavirus infections/statistics and numerical data”[Mesh]) AND (anxiety OR anxiety symptoms O anxiety disorders O anxious O “Trauma-related disorders and stressors” [Mesh] O “Anxiety” [Mesh] O “Anxiety disorders” [Mesh] O “Anxiety/epidemiology” [Mesh] O “Anxiety/statistics and numerical data” [Mesh]). No language restriction was made. This search was updated on August 31, 2022.

### Selection criteria

Inclusion criteria: studies that reported and/or related the existence of anxiety generated by suffering or not from diseases caused by the new emerging Covid-19 viruses and that for which the full text of the article was accessible.

Exclusion criteria: systematic or narrative reviews and studies in groups other than children and adults, examples of studies of communities, students, health personnel, population with associated comorbidities.

## RESULT

140 studies were identified from the initial search [figure 1 PRISMA flowchart]. After removing 11 duplicate studies, which were found in both Pubmed and Embase databases, 129 studies were examined; 86 of them were excluded because the full text was not available or they were systematic reviews, for which 43 full text articles were selected and evaluated for eligibility. Only 22 were considered for the qualitative synthesis and 21 additional full-text studies were excluded because they were not within the included time and were studies of population groups not selected for the review.

**Figure 1.**
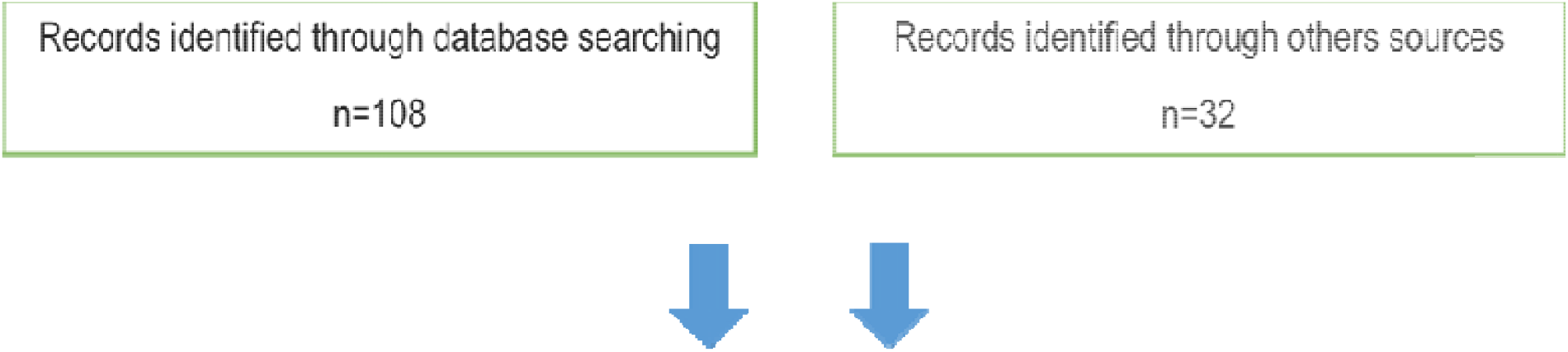
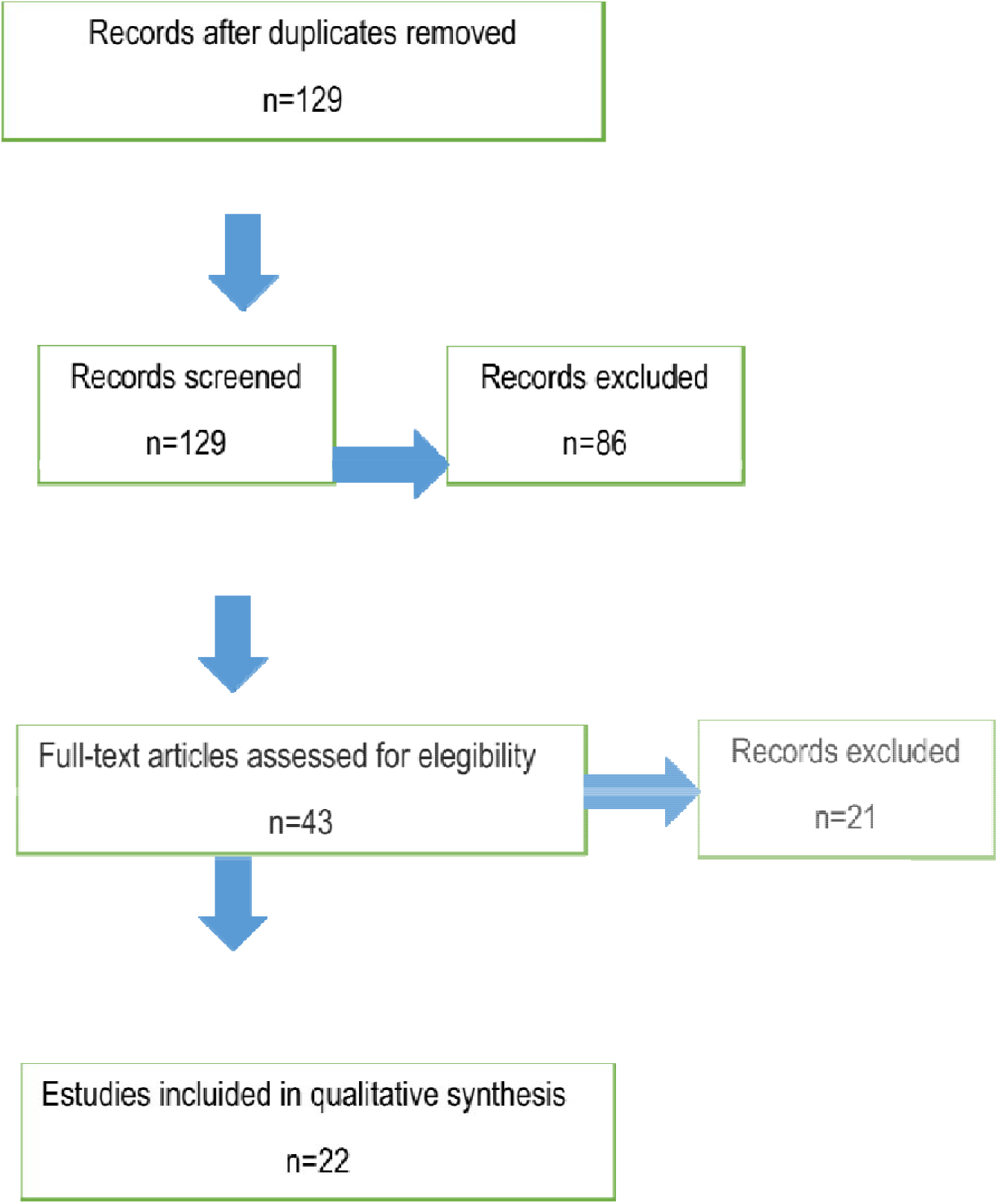
PRISMA flowchart.

In the 22 studies [see table 1: Characteristics of the included studies] selected for the qualitative synthesis, it was found that the main psychosocial repercussions were:

**Table 1.** Characteristics of the included studies.

| Studies | Characteristics |  |  |
| --- | --- | --- | --- |
|  | Design | Population | Results |
| Généreux M, et al. [22]. | Cross-sectional study design with a randomly selected, | Adult population (over 18 years of | Several sociodemographic, psychological, and sociocultural factors |
|  | interdisciplinary and international sample. | age), May to June 2020. | were found to be positively associated with a probable Generalized Anxiety Disorder or Major Depressive Episode, including (in descending order) weak SOC, younger age, false beliefs, isolation, perceived threat to self or family, mistrust of authorities, being a victim of stigma, threat perceived by the country or the world, financial losses, being a woman, and a high level of information about COVID-19.<br>Some of the factors found in this study to be associated with decreased psychological health in times of the<br>pandemic were female sex, younger age, discrimination, isolation, and less ability to adapt. |
| Idoiaga N, et al. [23] . | Descriptive study design. | 228 children aged 3 to 12 years, March 2020. | Children are afraid and worried about catching the virus, but mainly because they think they can infect their grandparents, and this makes them feel guilty. In addition, the confinement situation has produced mixed emotions in children. On the one hand, they are scared, nervous, lonely, sad, bored and angry. |
| Mactavish A,et al. [24] . | Observational, descriptive study design. | Children (ages 8-13) and their parents, in the Windsor-Essex region of southwestern Ontario. | Children and parents reported worse well-being and psychological distress during the pandemic. |
| Quero Acosta L, et al. [25] . | Observational, descriptive, multicenter (4 recruitment centers), and cross-sectional, random sample. | 150 individuals: 77 girls (51.3%) and 73 boys (48.7%) with a mean age of 10 .85 +/- 2.01 years. September to November 2020. | Reported psychophysiological symptoms of depression, anxiety and irritability. |
| Yasmin F, et al. [26] . | Cross-sectional study design. | adult population, March to July 2021. | The population with moderate to high psychological distress and low mental well-being, it can be stated that almost three quarters of the population in Pakistan under study had difficulties in adapting to the new state of life during the pandemic lockdown. |
| Khatatbeh M, et al. [27] . | Cross-sectional study design. | adult population, June to | a significant level of psychological burden on Jordanians, especially |
|  |  | September 2020. | women. |
| Qin Z, et al. [28] . | Cross-sectional study design. | child population, March 2020. | self-reported psychological distress. |
| Bala S, et al. [29] | Cross-sectional study design. | adult population aged 20 to 55 years, May 2020. | A significant positive association was observed for COVID-19 stressors, such as worry about the economy, the effect of social networks on daily life, poor knowledge, and a negative correlation of lack of social support with a higher score. IES-R in our study. The mean IES-R score was found to be at a higher end among low-resilience copers. |
| Ngoc Cong Duong K, et al. [30] | Cross-sectional study design. | adult population over 18 years of age, April 2020. | Respondents experienced psychological distress, depression, stress, and anxiety. Large-scale strict social distancing substantially increased anxiety, depression, loneliness, and substance use. |
| Wang C, et al. [31] | Cross-sectional study design. | adult and adolescent population. | Thailand had all the highest scores for stress, anxiety, and depression, and Vietnam had all the lowest scores. Risk factors for adverse mental health during the COVID-19 pandemic include age <30 years, high educational level, single and separated status, discrimination by other countries, and contact with people with COVID-19. Protective factors for mental health include male gender, being with children or more than 6 people in the same household, employment, trust in doctors, high perceived probability of survival, and spending less time on health information. |
| Traunmüller<br>C, et al. [32] | Cross-sectional study<br>design. | adult<br>population,<br>March to April<br>2020. | they rated the psychological<br>impact as moderate or<br>severe and reported<br>moderate to severe<br>depression, anxiety, and<br>moderate to severe stress.<br>Being a woman, older age,<br>lower educational level,<br>concern for family<br>members, Internet as the<br>main source of information,<br>status as a student or pupil,<br>self-perceived poor health,<br>and downplaying the<br>seriousness of the problem<br>were significantly<br>associated with a higher<br>psychological burden. |
| Passos L, et<br>al. [33] | Cross-sectional study<br>design. | adult<br>population,<br>March 2020. | Three weeks after the first<br>confirmed cases in<br>Portugal, participants rated<br>the psychological impact of<br>the outbreak as moderate or severe. In addition, they reported moderate to severe symptoms of depression, anxiety, and stress levels. |
| Tee ML, et al. [34] . | Cross-sectional study design. | adult population, March to April 2020. | psychological impact of the pandemic with higher levels of stress, anxiety, and depression. |
| Al Dhaheri AS, et al. [35] . | Cross-sectional study design. | adult population, May and June 2020. | Most of the participants felt horrified, apprehensive or helpless due to COVID-19. More than 40% reported increased stress from work and financial matters. Greater psychological impact was found among women, participants aged 26-35, those with a lower educational level, and participants residing in the North African region. |
| Barros MBA, et al. | Cross-sectional study design. | adults; April to | Frequent sadness and |
| [36] . |  | May 2020. | nervousness, as well as changes in sleep patterns, these mental health effects were greater in young adults, women and people with a history of depression. |
| Wang C, et al. [37] . | Longitudinal study design. | surveyed the general population twice: during the initial outbreak and the peak of the epidemic and four weeks later. | During the initial phase and four weeks later during the COVID-19 epidemic in China, there was a statistical but not clinically significant reduction in psychological impact. There were no significant temporal changes in levels of stress, anxiety, and depression between the start of the pandemic and four weeks later. |
| Alkhamees AA, et al. [38] . | Cross-sectional design. | population: adults between 30 and 40 years old. | A quarter of the general population experienced moderate to severe psychological impact. |
| <u>Davico</u> CH, et al. [39] . | Cross-sectional study. | children and adults in March 2020. 2,419 adults and 786 children. | Serious psychological impact and high risk of post-traumatic stress disorder in children were observed. Children may also be more vulnerable due to home confinement, school closures, lack of personal contact with classmates, friends, romantic partners, and teachers, and limited personal space at home. |
| Saddik B, et al. [40] . | cross-sectional study. | adults and children during March to May 2020. | high anxiety of parents and children. |
| Thasneem Zainudeen Z, et al. [41] . | Cross-sectional study. | 409 parents and 348 children | 154 respondents (38%) reported high psychological impact (score 14) for the psychological construct, and 189 respondents (46%) reported high psychological impact (score 6) for behavioral construct. A<br>significantly higher proportion of respondents with non-permanent employment status of the head of the family reported high psychological impact. The prevalence of anxiety reported by the family members surveyed was 23%. Forty-five children answered the DASS-21 questionnaire; 28.5% reported anxiety, 31.4% depression and 13.3% stress. The family leader's job security status was found to be the predictive factor for mean total IES-R score (psychological construct) and ethnicity for mean total CRIES-8 and CRIES-13. Conclusion Rates of depression and anxiety during the COVID-19 pandemic were high. |
| Gutiérrez<br>Álvarez AK,<br>et al. [42] . | Descriptive design,<br>corresponding to the<br>first stage of spread<br>of the disease in the<br>country. | Population<br>children and<br>adults. | The time of social isolation caused the restructuring of the lives of its inhabitants, which generated readjustments in both family and community roles and functions.<br>The main impact on the community was associated with socioeconomic factors; It manifested itself fundamentally in the difficulty of having access to basic consumer goods.<br>Decompensation of comorbidities due to the stress generated.<br>The natural processes of resilience of the community were developed, which were fostered by a government management that gradually gained order and efficiency. |
| Herrera Paz | Observational study. | Adult | sadness, depression, |
| JJ, et al. [43] |  | population. | anxiety, financial concern,<br>fear of contagion of COVID<br>19. |

- restructuring of life with readjustments in both family and community roles and functions.
- establishment of unhealthy habits (eg poor eating habits, irregular sleep patterns, sedentary lifestyle and greater use of technological means) with derivation of physical problems.
- increase in depressive and anxiety disorders where people’s mobility was lower and the daily rates of COVID-19 infection were higher.
- mass hysteria, financial burden and financial loss. The massive fear of COVID-19, called “corona phobia”.
- affectation in frequency, sharing, communication, closeness and emotional support in their relationships with family, friends and neighbors or other people, mostly experiencing emotions such as sadness, stress, fear and anxiety.
- rejection or discrimination for having presented the disease, uncertainty of the prognosis of the disease, misinformation about the disease, financial insecurity.
- The fear of infecting vulnerable people and non-compliance with academic and work demands caused frustration, worry, fear, disgust, nostalgia and guilt, thus concluding that the participants experience mild symptoms of anxiety and stress, in addition it was shown that the social support is a protective factor that favors the attenuation of symptoms.

The circulating variants responsible for these main psychosocial repercussions were: *Epsilon, Zeta, Eta, Iota, Kappa, Alpha, Beta, Gamma, Delta. Alpha, Beta, Gamma* and Delta variants were distributed throughout the world and were responsible for a second and third wave of the pandemic. The first wave predominantly infected older and immunocompromised populations and people with underlying complications. In the second wave of the pandemic, an Alpha variant that mostly affected the young population with community transmission and hospitalization, and the Gamma variant responsible for reinfection in the state of São Paulo, Brazil, were reported. The Delta strain drove the second wave in India and was responsible for the third wave in the UK. The limitations found in the articles reviewed were: mostly cross-sectional studies were carried out and online surveys were generally used to obtain information. The studies are mainly in the adult population and when they are carried out in the child population they are accompanied by the participation of adults (parents). No studies were found that evaluated the psychosocial impact in correspondence with each variant of COVID 19.

## DISCUSSION

Our review found that, during the pandemic, there was an alteration of mental health with a very large psychological impact on all age groups where anxiety, stress, different levels of depression and difficulties in adapting to the surrounding environment prevailed. COVID-19 blockages have become a major psychological risk, which poses a challenge in times of different pandemics, especially in children and adolescents. [44]

In children, peaks of anxiety and irritability could be observed and they were aware of the psychological distress it caused them. Factors that were associated with vulnerability were confinement, school closures, lack of contact with other people and limitations of personal space. Interestingly, among the factors influencing the children was the fear they had of becoming infected and infecting their grandparents, who reported feeling scared, guilty, lonely, sad and angry. Another study showed that children with psychiatric and/or developmental problems were more affected psychologically; family and social support was recommended as a preventive strategy. [45]

Among the results, it was found that the main factors associated with psychological impact were being female, average age between 26 and 35 years, low level of education, being single, difficulty in accessing basic consumer goods, sadness, personal worries, fear of contagion, having a previous history of depression; in addition, among those associated with the adult population were not having a permanent job, family worries and using the internet as a source of information. This is in accordance to results found in previous studies. [46-49]

On the other hand, the factors that were found to be protective were being male, living in houses with children or more than 6 people, stable employment and spending less time on health information. This is similar to what has been found in studies and reviews already carried out where the company becomes an important point in the way of coping with the psychological impact of the pandemic; [50] therefore, promoting and guiding the obtaining of mental health should be considered a public health priority.

## Data Availability

All data produced in the present work are contained in the manuscript

## FUNDING

No extramural funding was received.

## COMPETING INTEREST

This is to declare that the authors do not have any conflict of interest.

## ACKNOWLEDGEMENTS

The authors would like to acknowledge the open access resources from where data was collected under Creative Common License.

## Notes

### Competing Interest Statement

The authors have declared no competing interest.

### Funding Statement

This study did not receive any funding

### Summary of Updates

This version of manuscript was revised to update the title page

